# Rapid Lifestyle Prototyping: Assessment of Infrared Spectroscopic Blood Responses for Short-term Lifestyle Interventions

**DOI:** 10.1101/2021.08.07.21261731

**Authors:** Dara Rouholiman, Erik Andries, Sylvie D Dobrota, Hershel Macaulay, Robert G. Messerschmidt, Thomas Quertermous

## Abstract

**Background:** At the present time, optimization of dietary and fitness practices are inadequately addressed in the clinical setting, and poorly documented through research studies investigating response to surrogate disease markers. Identifying and quantifying the personal response to lifestyle interventions requires a new tool and a new measurement metric.

**Methods:** In this prospective observational study, COR.RELATE study, we investigated the effect of four lifestyle interventions, each spanning 21 days and consisting of seven validated practices, in a healthy population. We used a novel at-home near-infrared spectrometer (wavelength 1600-2450 nm), COR console, to measure the spectral blood changes in a cohort of 178 healthy males and females who participated in four 21-day cycles of lifestyle programs. In the first round, study participants also completed dried blood spot cards, provided by omega quant, at the start and end of their program.

**Results:** We analyzed 1,870 infrared spectroscopy samples from 178 participants (131 males and 47 females) who participated in the first four programs of COR.RELATE. We found on average 79.75% of the participants are responsive to one or more of the COR programs. We describe an algorithmic approach, ranking personal responses to different nutrition and fitness interventions.

**Conclusion:** The COR console and data analytics enables the use of a novel near-infrared based metric--which we call the COR Blood Response Pattern (BRP)--to measure individual response to lifestyle change, and has the sensitivity to discern, classify, and rank the success of rapid lifestyle interventions in terms of the correlated spectroscopic response they produce. Rapid Lifestyle Prototyping is a new concept in consumer life optimization and improvement - the ability to know in no more than 21 days whether a particular single, or set of, lifestyle practices is going to evoke a strong response for an individual and therefore if it is worth continuing that practice in their quest for deep optimization and life improvement.

## Introduction

Randomized clinical trials (RCTs) are considered the gold standard in both clinical and nutrition and fitness research. RCTs evaluate the association between nonrandomized effect and outcomes and provide scientific evidence on the effect of an intervention.^1^ However, RCTs are limited when it comes to comparing different nutrition and fitness interventions.^2-4^ There is a growing need for individuals searching to rapidly optimize their healthy lifestyle with a tool and a metric to compare and contrast different nutrition and fitness interventions in a period of time short enough to get and maintain engagement.

Existing nutritional and activity guidelines are not one size fits all, as the variance between groups and individual responses to lifestyle interventions is often not highlighted in traditional clinical studies, with conclusions and recommendations being based on the average response of the study populations. ^3,4^ Results from recent studies suggest that an individual’s biochemical response to lifestyle interventions is unique, and may not reflect the population average. The PREDICT study enrolled a large cohort of twins and unrelated healthy adults (n=1,002) and found large inter-individual variability in postprandial responses to identical meals. ^5^ In addition to large heterogeneity in the population’s response to nutritional and fitness interventions, the studies and the tools used to measure these responses have severe limitations. Studies often suffer from high inter-individual variability in compliance to lifestyle interventions. ^5,6^ Further, the allowable analytical error margins in for surrogate disease markers such as lipid and lipoprotein levels are large, as high as 30% from the target value for some measurements.^7,8^ This margin of error is much larger than the 5-10% change in concentration of most individual biomarkers in the blood seen after several weeks of doing an intervention. ^9-17^ These existing limitations have motivated us to design a novel infrastructure and analytical methodology to identify and quantify with high precision the subtle molecular changes in the blood that result from exercise and dietary changes.

In the COR.RELATE study, we investigated the use of an at-home near-infrared spectrometer, the COR console, to detect ensemble spectral changes which can be due to chemical, structural, and environmental changes in blood samples taken from healthy adults living in the United States. The COR console uses optical technology, molecular spectroscopy, to measure whole blood and blood plasma spectra, in a very small capillary blood sample. The precision of the COR console allowed us to reproducibly assess spectral changes in blood molecular patterns that are largely undetectable with other tools. We analyzed the association between the spectral changes and lifestyle programs.

Our findings show that common-mode spectral changes of a population have large variance and are attributed and correlated in large part to prescribed programs. We found that practices in the programs each have different magnitudes of effect on the spectral changes and people have different responses to different programs. The instrument and the BRP metric we have developed could be used to better optimize personalized lifestyle recommendations for discovering strategies to ultimately improve individual health.

## Methods

### Study Design Overview

COR.RELATE was a double arm study consisting of seven total COR Programs, each lasting 21 days (Figure 1). COR Programs contained a combination of lifestyle tasks of exercise, attention, recovery, and nutrition (EARN) focused tasks.

**Figure 1:**
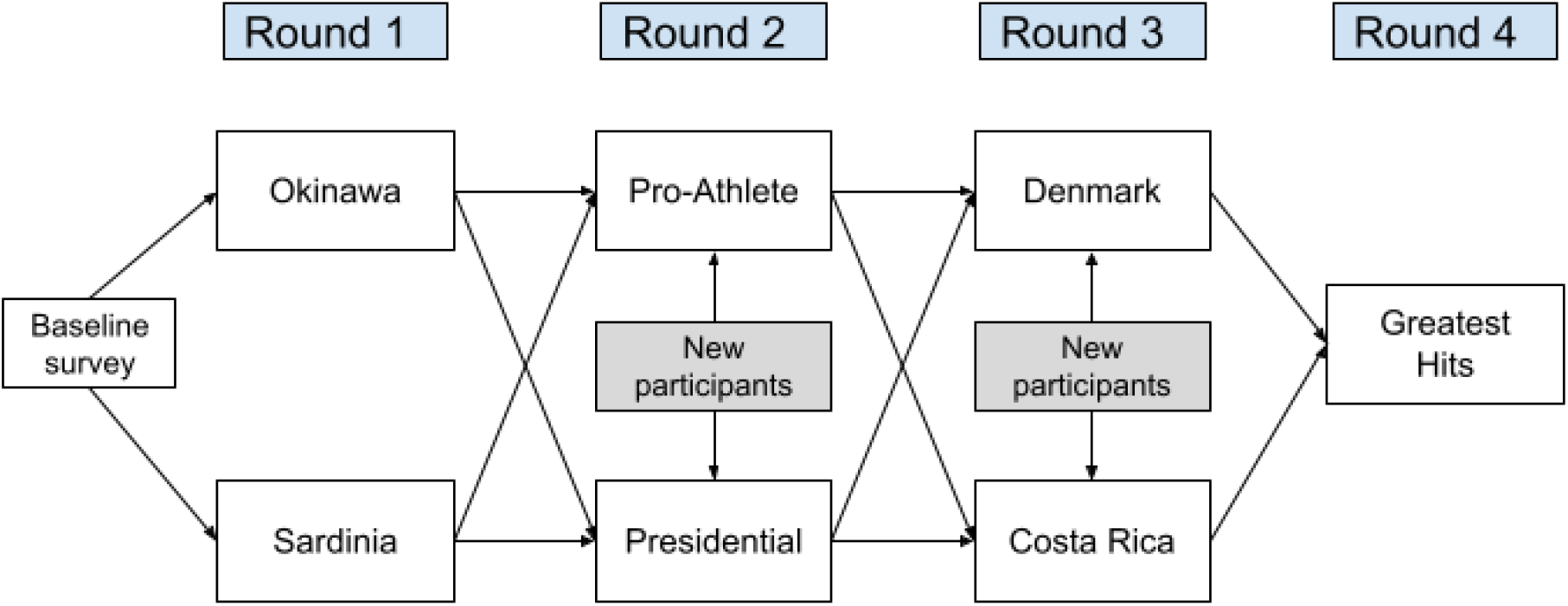
The COR.RELATE double-arm study design- Participants selected one of two programs to complete for each of the first three rounds.

Participants were able to choose between two COR Programs at the beginning of each round of testing. During each COR Program, and throughout the study, participants completed regular blood samples assessments on the COR console, called COR check-ins. In the first round, study participants also completed dried blood spot cards, provided by OmegaQuant, at the start and end of their program. These cards were analyzed by OmegaQuant for Omega-3 index, Trans-fat index, Omega6:Omega3, and AA:EPA.

### Participants

All potential participants were screened for eligibility by answering a few short questions online. We were interested in individuals that:

a. Showed a high interest in optimizing their health and well-being
b. Were open to conducting and testing new nutrition & fitness routines
c. Met eligibility criteria

Individuals were eligible if they were generally healthy and between 21 and 70 years of age. Full inclusion and exclusion criteria are listed in Supplemental Table 1. Participants that met eligibility criteria were loaned a COR console, cartridge and lancets for home use after signing the informed consent.

### COR Console

The discovery of near-infrared energy is attributed to William Herschel in 1800,^18^ was first applied as a quantitative tool in the field of agriculture, led by the work of Karl Norris,^19^ and has been successfully applied to biochemistry and medical applications since at least 1977^20^ The most well known application of near infrared spectroscopy, albeit at much shorter wavelengths) is almost certainly that of pulse oximetry, in use almost constantly in clinical practice today. More recently infrared spectroscopy has been applied in researching single cells.^21^

The COR console is a spectrometer with the wavelength range of 1600-2450 nm (resolution of 7-8 nm). It captures the NIR spectra of the blood in three stages, including for whole blood, in-transition, and plasma as blood gravimetrically separates over a time period of 90 minutes. The spectrometer employs a number of improvements over previous generation instruments. The focus of the improvements is on providing high repeatability of sample insertion, temporal resolution, high thermal stability, high SNR, and a consumer level price point. Various wavelength regions in the near-infrared were considered and this was found to be the best. The spectrometer operates at the very longest wavelength end of the range classically considered near infrared; this region of the infrared spectrum contains voluminous molecular information.

### COR Programs

Each COR Program consists of the fundamental elements and defining characteristics of an inspiring health culture, designed for routine daily practice over a three-week period. Each COR Program includes nutrition, fitness, and recovery ideas on which to focus. Subjects were instructed to try to complete each of seven items in a series, in order, every day, and check them off on the app as they make progress through the list as shown in Figure 2.

**Figure 2.**
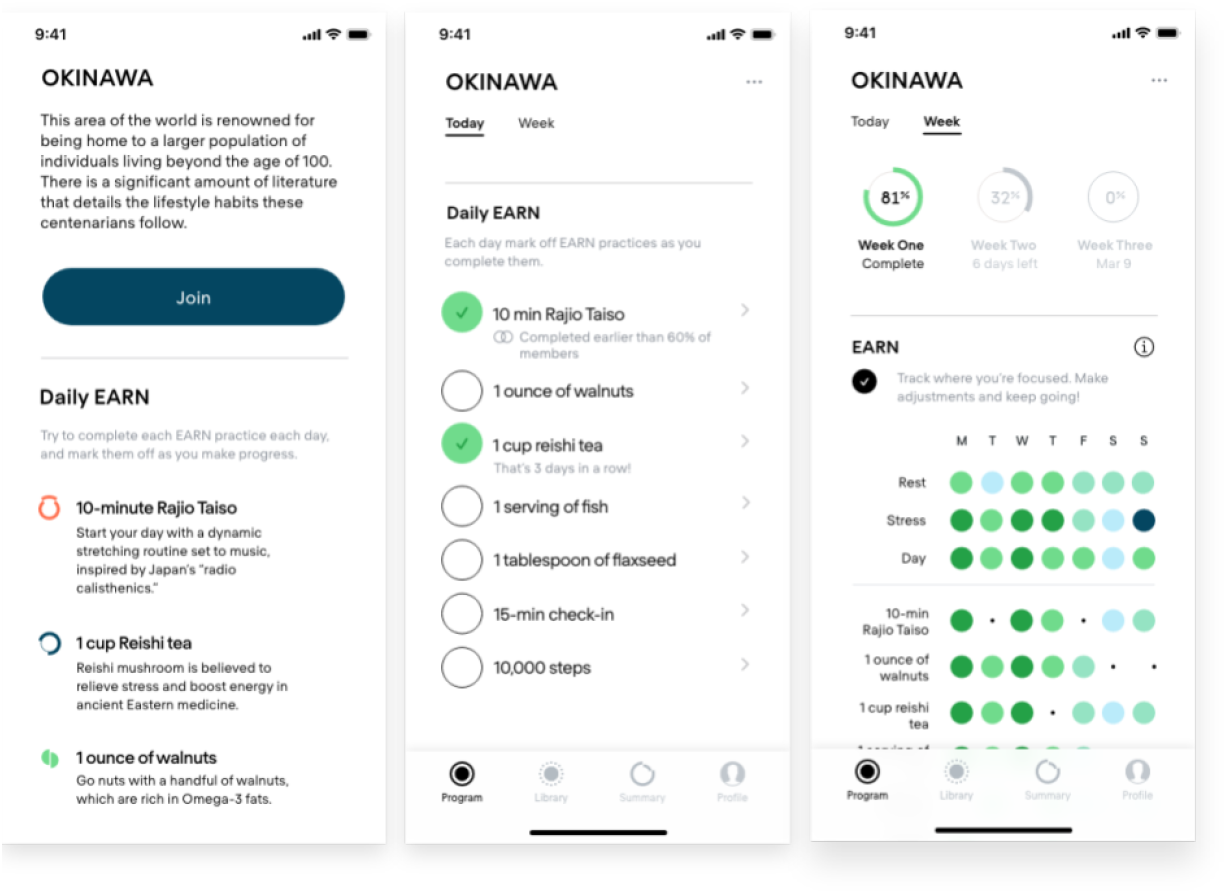
App Screenshots-Participants joined a program in the app (left) and marked off the practices they completed daily over the course of 21 days (middle). Participants could access a weekly view of their practices (right).

### Data analysis

To find and measure the magnitude of change in blood due to the set of lifestyle practices, we measured the overarching, subtle infrared pattern changes in blood in each program and assigned a statistically significant pattern change occurring across that population that is due to the process of completing the practices of the program. We call these patterns blood response patterns (BRPs) as a measure of magnitude of change in blood due to the unique pattern that arises from an ensemble of all molecular changes in the blood that correlate with the program activities, measured in a score from 20-100.

The aim of the data analytics is to characterize the portion of the total blood response that is indeed correlated statistically and across the population with the doing of the lifestyle practices. That way we can be confident that the BRP results from and is evoked by lifestyle practices that have been already validated as beneficial in the medical community and literature. Participants who did not complete at least two check-ins during the program or more than 30% of the practices were dropped from the analysis data set. Each COR program consisted of two or more COR check-ins, taken at least 2 days apart, to capture the BRP. A check-in captures absorption spectra of blood at three different physical states (whole blood, in-transition, plasma) over time. At each distinct blood state, a difference spectra between check-ins is used to capture the blood change as shown in Figure 3. The data structure of our analysis is a tensor for an absorbance with wavelength, blood state, and time measurements.

**Figure 3.**
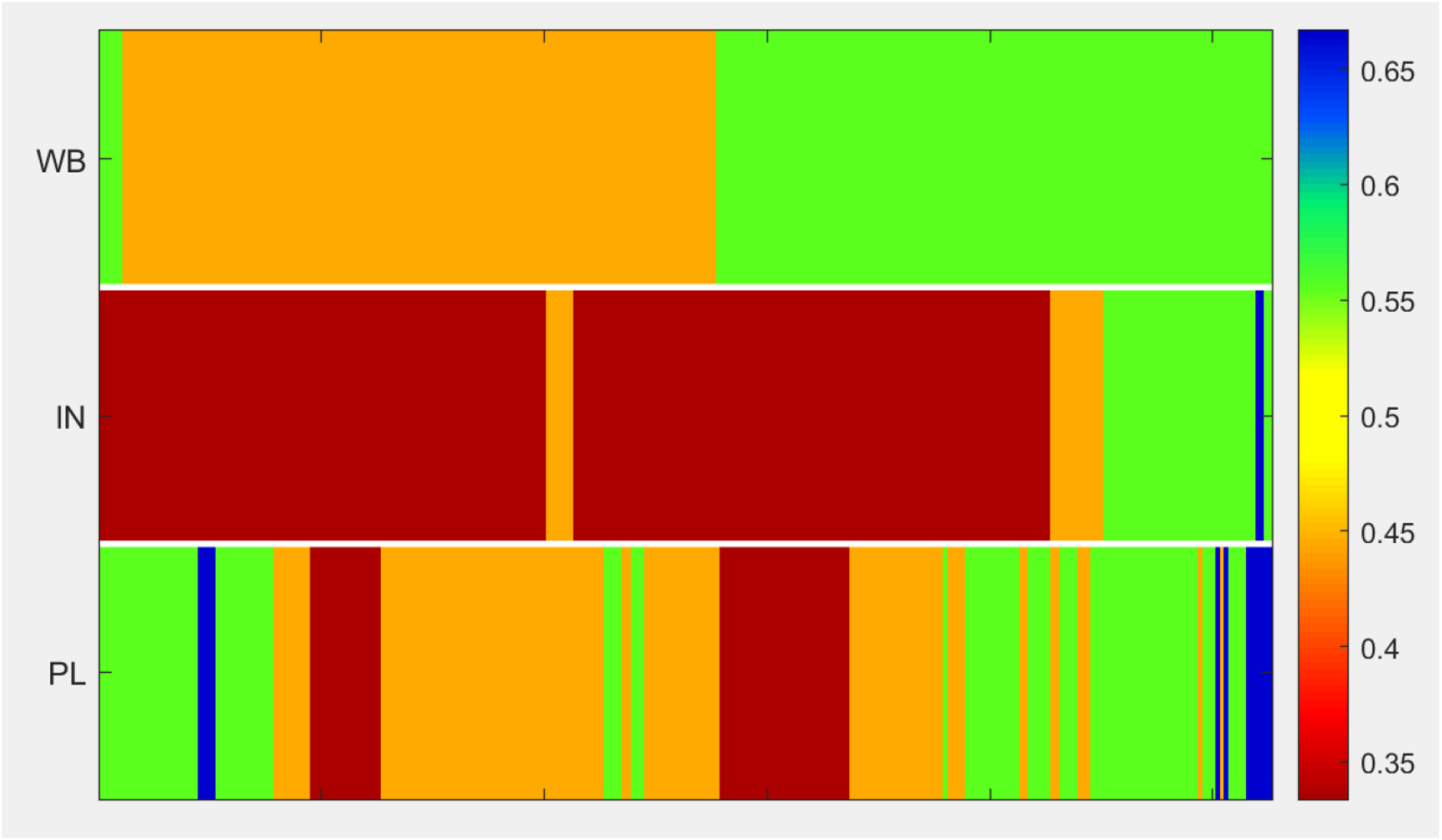
Percent Difference Spectral heatmap, a user’s spectral blood change in a 21 day COR program with 10 blood samples. The color shows the percent difference in absorbance at a wavelength in each blood state. Whole-blood(WB), In-transitions (IN), Plasma(PL).

## Results

263 healthy adults, aged 18-75, from the US, met eligibility criteria and were enrolled in the COR.RELATE study. Of these,178 individuals were included in the analysis of the first four programs. Table 1 presents characteristics of the participants at the baseline. The study design is described in the Methods and Figure 1, the inclusion and exclusion criteria is presented in Supplemental Table 1.

**Table 1.**
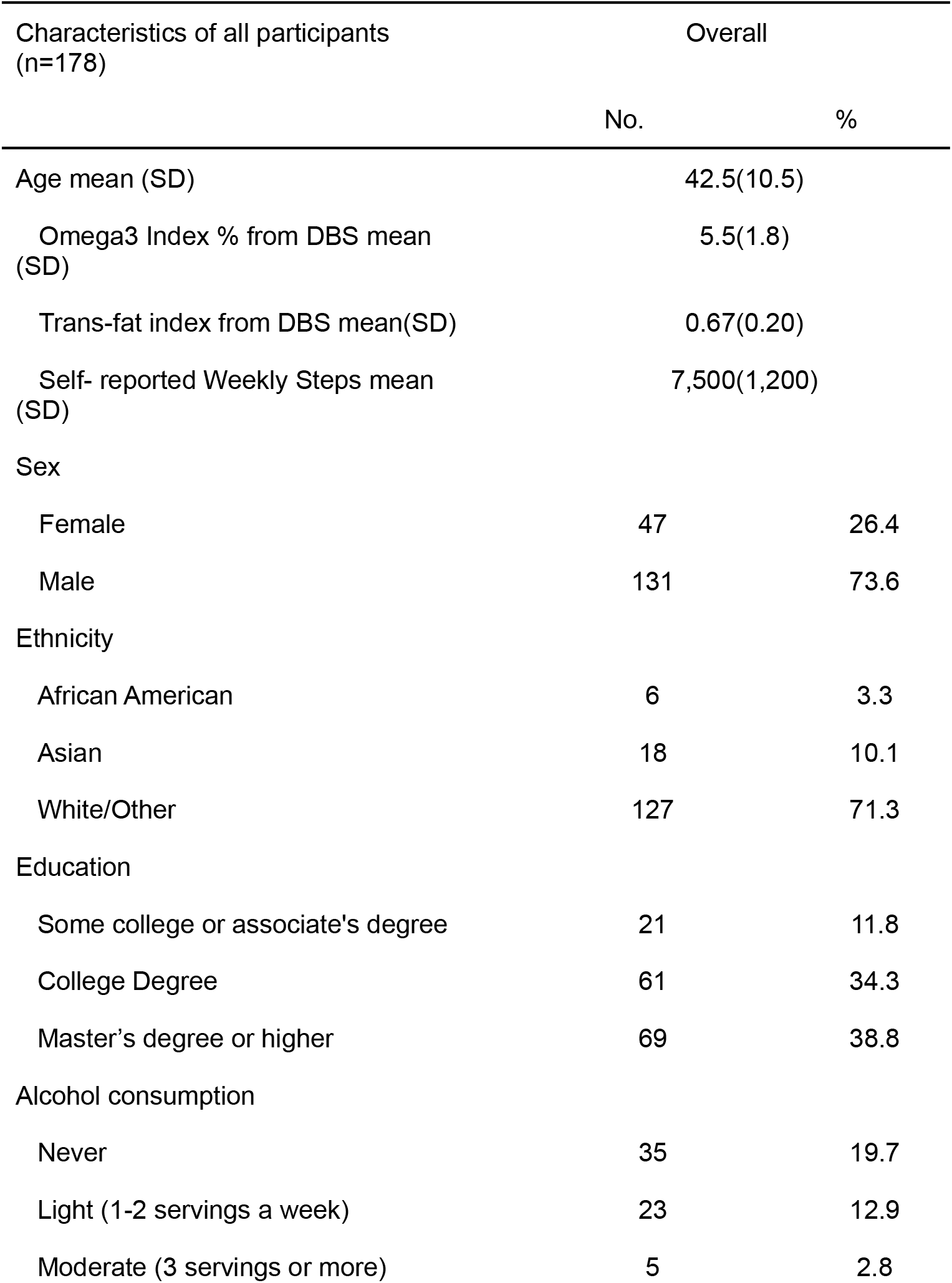
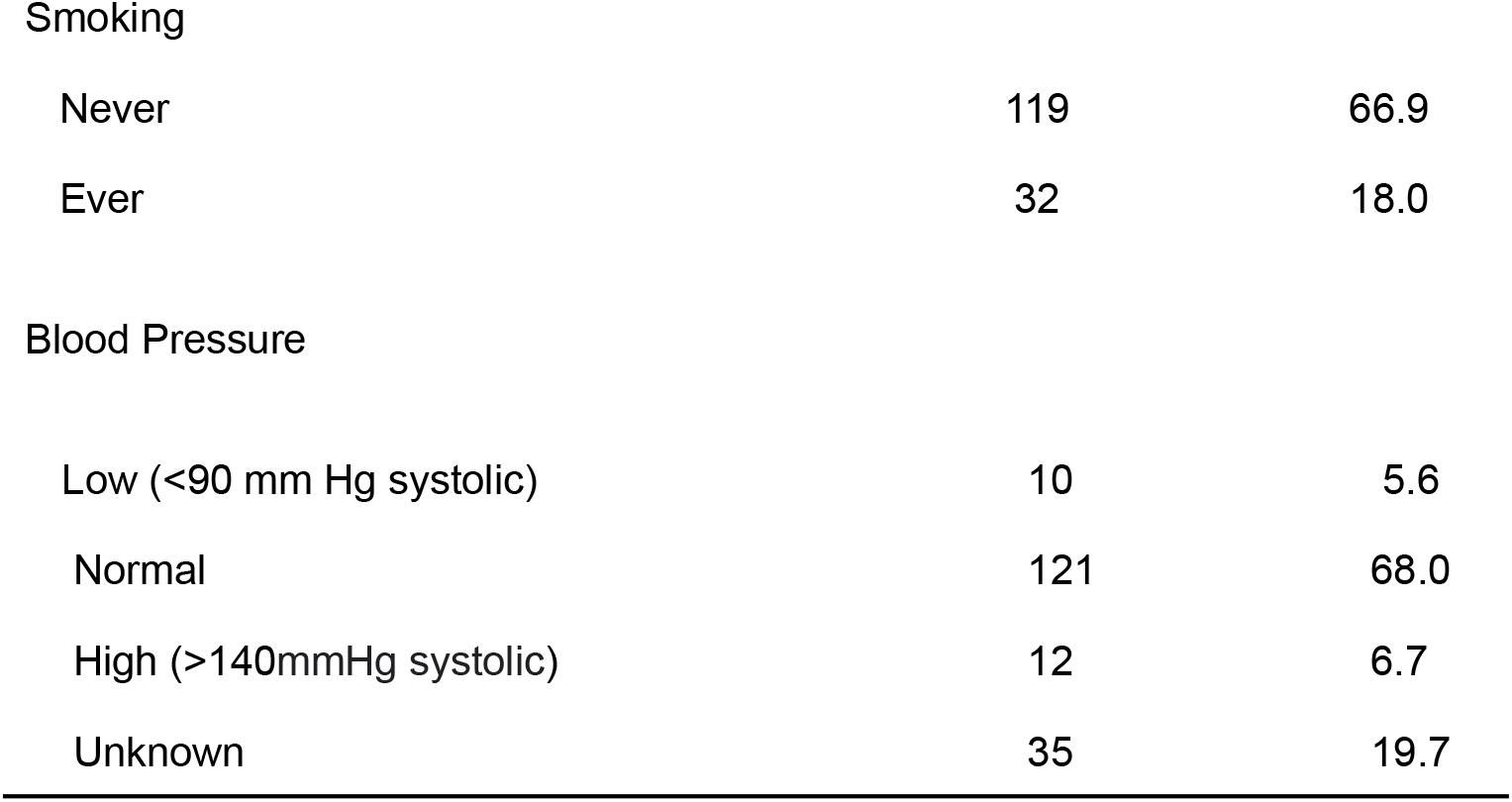
Characteristics of COR.RELATE participants.

| Characteristics of all participants<br>(n=178) | Overall |  |
| --- | --- | --- |
|  | No. | % |
| Age mean (SD) | 42.5(10.5) |  |
| Omega3 Index % from DBS mean<br>(SD) | 5.5(1.8) |  |
| Trans-fat index from DBS mean(SD) | 0.67(0.20) |  |
| Self- reported Weekly Steps mean<br>(SD) | 7,500(1,200) |  |
| Sex |  |  |
| Female | 47 | 26.4 |
| Male | 131 | 73.6 |
| Ethnicity |  |  |
| African American | 6 | 3.3 |
| Asian | 18 | 10.1 |
| White/Other | 127 | 71.3 |
| Education |  |  |
| Some college or associate's degree | 21 | 11.8 |
| College Degree | 61 | 34.3 |
| Master's degree or higher | 69 | 38.8 |
| Alcohol consumption |  |  |
| Never | 35 | 19.7 |
| Light (1-2 servings a week) | 23 | 12.9 |
| Moderate (3 servings or more) | 5 | 2.8 |
| Smoking |  |  |
| Never | 119 | 66.9 |
| Ever | 32 | 18.0 |
| Blood Pressure |  |  |
| Low (<90 mm Hg systolic) | 10 | 5.6 |
| Normal | 121 | 68.0 |
| High (>140mmHg systolic) | 12 | 6.7 |
| Unknown | 35 | 19.7 |

### User Engagement

While in the study, participants were instructed to do 10 COR check-ins (that is 10 finger prick blood samples) in each program and log their program practices in real-time via the COR app. On average participants were highly compliant with the average 6.8 COR check-ins and average daily practice of 5.2 practice out of 7 in the first four programs.

### Inter-individual variability in blood responses

The Blood Response Pattern response of participants in each program were measured using that program’s BRP with the median BRP responses of 53, 49, 48, and 52 for Presidential-Athlete, Sardinia, Pro-Athlete and Okinawa programs respectively (Figure 5a) The inter-individual patterns of response for each program was assessed using Levene’s test of variance.

**Figure 4.**
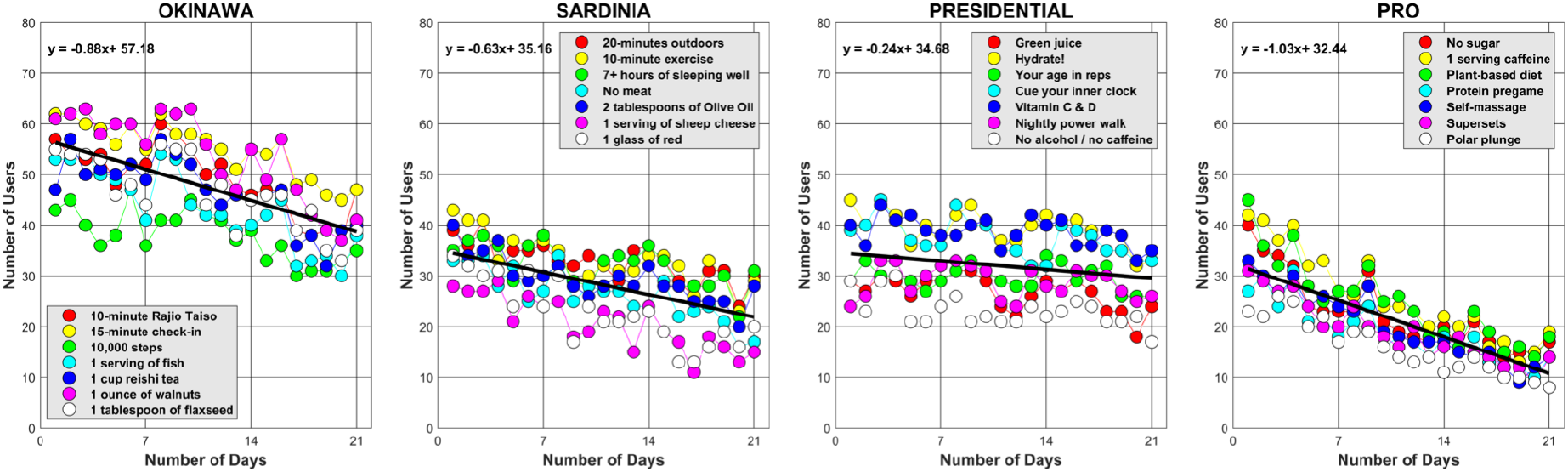
Users’ engagement with program practices in 21 days. In all four programs, the engagement is considerably decreasing after day fourteen.

**Figure 5:**
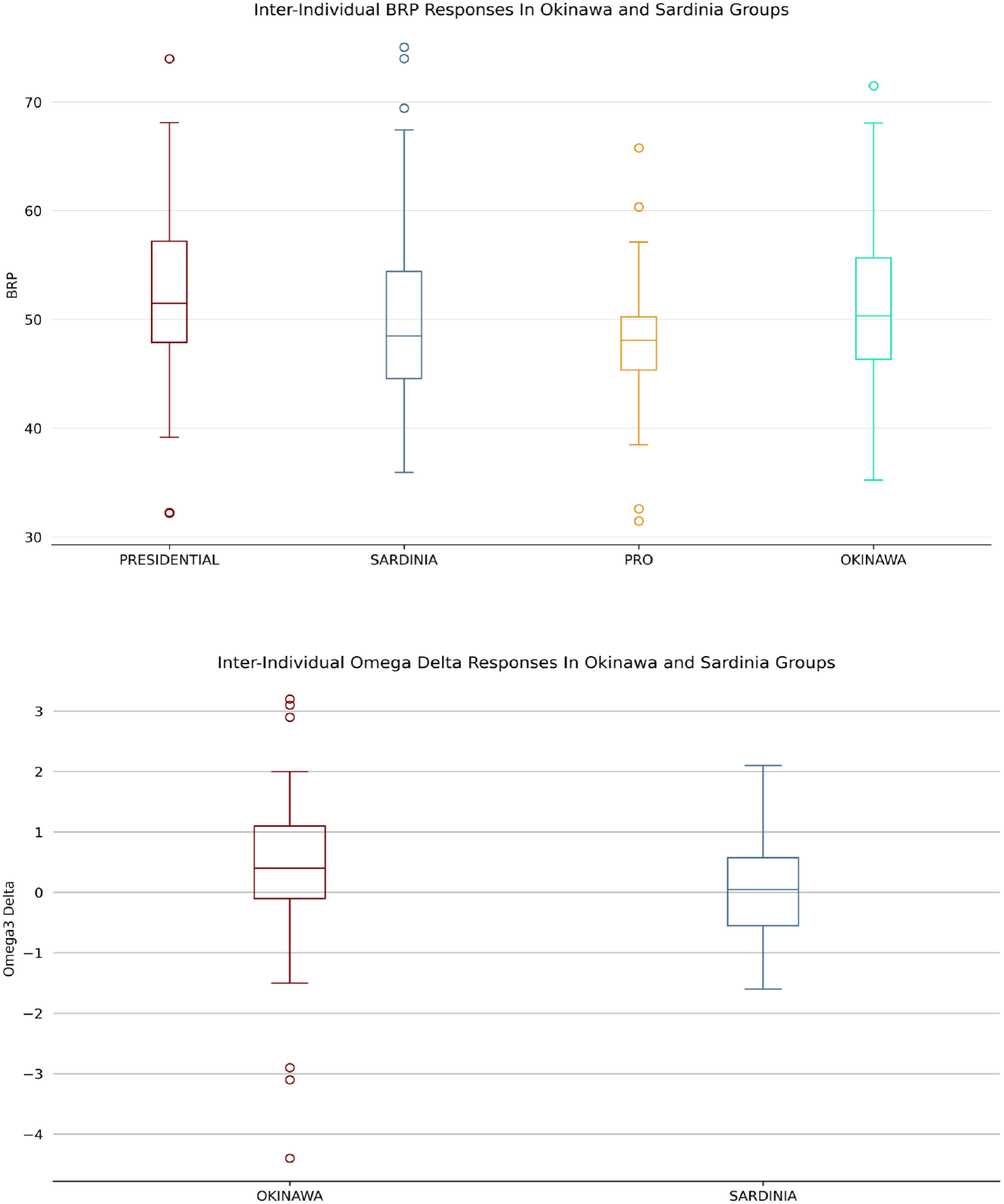
(top) Blood response pattern (BRP) responses in Okinawa and Sardinia participants. (bottom) Omega-3 index dried blood spot responses in Okinawa and Sardinia participants.

To characterize the expected change in blood Omegas, in Okinawa and Sardinia, participants were asked to do Omega Quant’s dried blood spot card one on day 0 of the program and another on day 21. In Figure 5b the omega3-index change(delta) in the two programs is shown.

Further, the BRP values were compared with the Okinawa’s engagement and its seven daily practices using Pearson correlation coefficients (r); these correlation were r=0.24 (p >0.1), r= 0.31(p=0.04), r=0.02 (p >0.1), r=0.12 (p>0.1), r=0.16 (p >0.1), r=0.16 (p >0.1), r=0.02 (p >0.1), r=-0.002 (p >0.1) for overall program activity, 10-minutes Rajio Taiso, 15-minute check-in, 10,000 steps, 1 serving of fish, 1 cup of reishi tea, 1 ounce of walnuts, and 1 tablespoon of flaxseed respectively. The heatmap of these correlations are shown in Figure 6.

**Figure 6:**
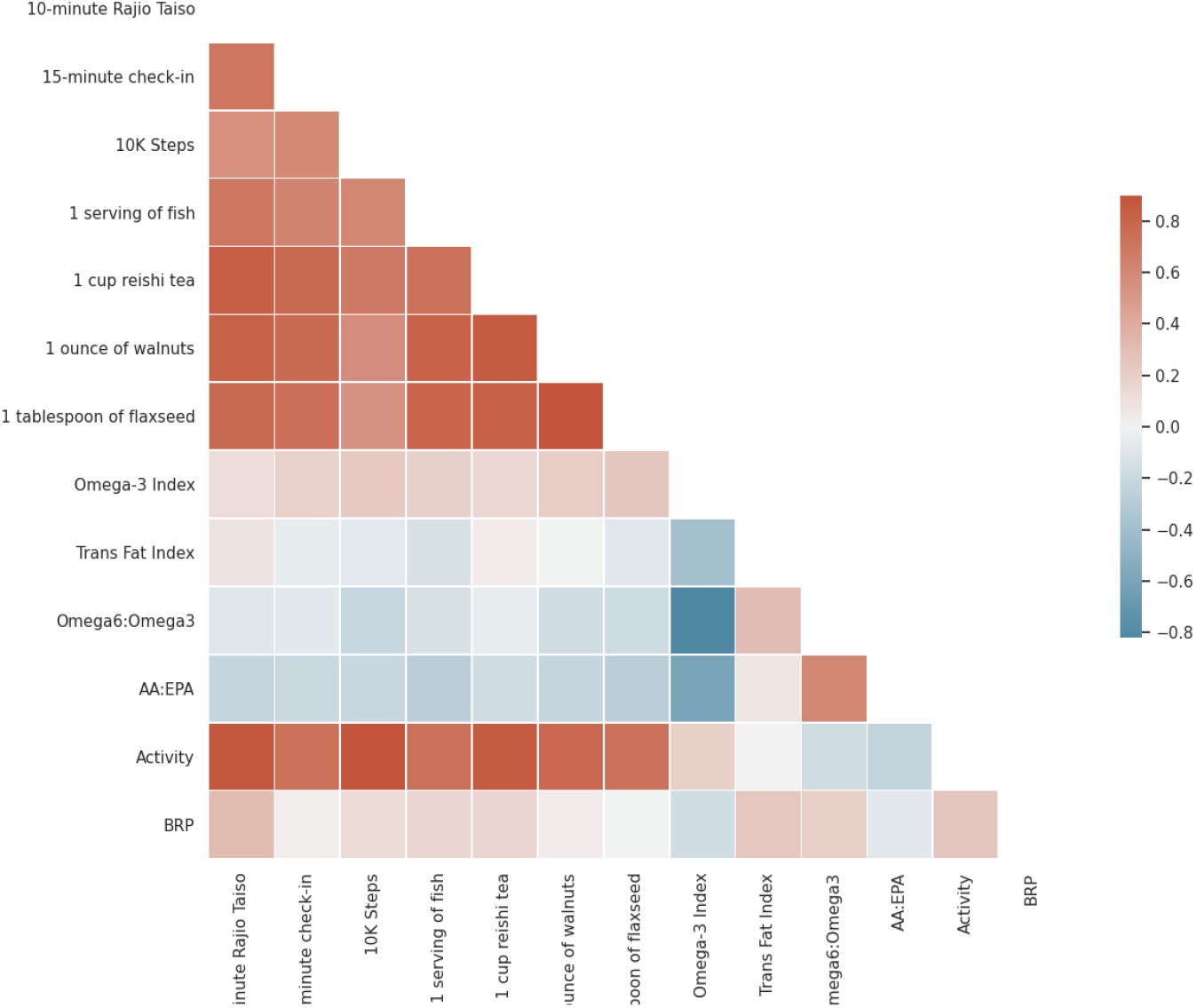
Heatmap of statistically significant correlations (Pearson) between BRP, Omega3-index delta and Practices engagements in Okinawa cohort.

**Figure 7:**
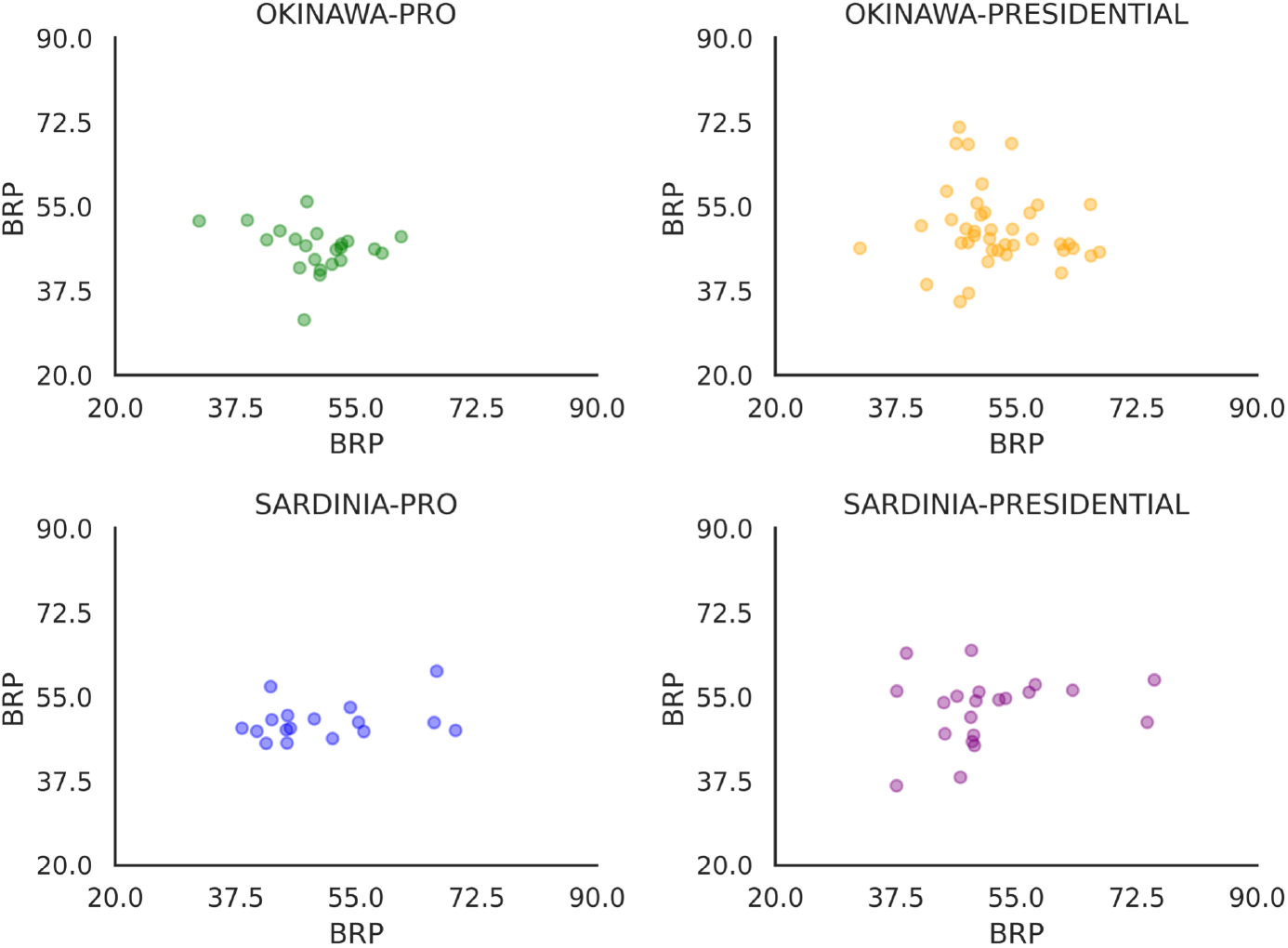
Blood response pattern (BRP) responses of individuals across two programs. (Top Left) BRP values of those completed both Okinawa and Pro-Athlete Programs, Their Okinawa BRP on the y-axis with respect to their Pro-Athlete BRP on the x-axis. (Top Right) BRP values of those who completed both Okinawa and Presidential Programs. (Bottom Left) BRP values of those who completed both Sardinia and Pro-Athlete Programs. (Bottom Right) BRP values of those who completed both Sardinia and Presidential-Athlete Programs.

### Intra-individual variability in BRP responses

Having investigated BRP responses within the study population, we then explored the responses at the individual level. On average 79% of participants were responsive(*BRP >=50*) to one or more programs (Table3). Finally we used the BRP of participants in two common programs to ask whether the interventions induced different BRP changes. Figure7 shows the relationship between the BRPs of participants in two programs.

**Table 3:** Individual responsiveness to the programs (responsive: BRP >=50, unresponsive: BRP< 50)

| Program | % Responsive |
| --- | --- |
| Okinawa | 88 |
| Sardinia | 67 |
| Presidential-Athlete | 84 |
| Pro-Athlete | 80 |

## Discussion

### User Engagement

Our Cohort is representative of the ultra healthy adult population in the US, a population that is expected to be highly motivated to optimize their nutrition and fitness. Our data (Figure 4) shows that our cohort’s engagement and adherence to their EARN practices start to significantly decrease after day fourteen. The engagement decreases after only two weeks while most intervention studies in the literature are 6-8 weeks long, underscores a need to have a sensor that can detect subtle changes in the blood during short interventions rather than a blood biomarker concentration test that is not expected to change significantly for weeks or months.

### Inter-individual variability in blood responses

Each COR program consists of seven lifestyle practices that are validated by others to have positive effects on one’s health. In the Okinawa program, eating salmon daily is a nutritional practice that is widely studied, known to increase Omega-3 concentration in the blood. Increase in blood Omega-3 has positive effects on lowering cardiovascular risks and improving longevity.^22^ The BRP of the Okinawa program has captured the effects of this practice in combination with other six known healthy practices on the blood. An individual’s BRP result can tell us about their responsiveness to the program-a BRP within or higher than the 50th percentile can be regarded as responsive and a number below the 50th percentile box is regarded as non-responsive to a program. The BRP is quantitative with a larger number indicating more response to the known-good healthy practices.

In line with studies that examined Salmon intake and Omega3, the Omega3-index from the dried-blood spot cards did show an increase for the Okinawa cohort, although not as big of an increase as other studies. No Omega3 change was detected in the Sardinia cohort. We did not use the omega data in our BRP analytics. We used omega dried-blood spots to confirm our hypothesis that subtle changes do happen in short 21-days interventions. As expected, the program with a daily fish diet shows a change in Omega3-Index and the program with no omega3 intervention does not.. The Omega3-index and BRPs did not show any association, although BRPs correlated with program engagement.

### Intra-individual variability in BRP responses

One of our worries in the study was the influence of subject bias on the BRP models. In particular, the highly responsive participants would be the same participants across programs, and likewise for the low-responsive participants. We studied the relationship between BRP variables of the same participants completing the same programs and found no association between them. This result suggests that there is not simply one program that is most impactful for everyone; participation in COR program sequence has the ability to match individuals with the particular programs that are most impactful for them.

## Conclusion

We built a new blood analysis spectrometer-based system that is highly sensitive and with high precision, not to individual biomarkers, but to “ensemble” spectroscopic change. This system allows assessment of individual responses to exercise and dietary fitness modifications.

However, given that nutrition and fitness interventions vary in response in different people, this would not be possible without employing a “forcing function” to evoke objectively positive changes in blood response spectra. This approach characterizes the magnitude of response across a user population and indicates whether an individual response is consistent with the expected change in the blood response pattern. If so, a subject would be expected to benefit from continuing that particular COR program.

We used hundreds of COR spectrometers to conduct IRB-approved in-home human studies of blood response using well accepted clinically validated lifestyle interventions. Using only the COR spectroscopic data, we found spectroscopic patterns that correlated with programs and practices. We then ranged and ranked the magnitude of this correlated response we observed amongst a cohort of study participants so that we can identify the individual response and report this information to the participant.

We have demonstrated that the COR console and data analytics have the sensitivity to discern, classify, and rank lifestyle interventions in terms of the correlated blood spectroscopic response they produce. We can be sure that the blood response is beneficial by always starting with the most highly researched, most highly validated lifestyle interventions known to biomedical science. In this manner, we have created a new concept in consumer digital health which we call Rapid Lifestyle Prototyping - the ability to know in no more than 21 days whether a particular single, or set of, lifestyle practices is going to evoke a strong response for you as an individual and therefore if it is worth continuing that practice in a quest for deep optimization and life improvement.

## Data Availability

The data that support the findings of this study are available on request from the corresponding author, [TQ]. The data are not publicly available due to [restrictions e.g. their containing information that could compromise the privacy of research participants

## Supplemental Tables and Figures

**Supplemental Table 1.** Inclusion and exclusion criteria

| Subject Inclusion Criteria: | Subject Exclusion Criteria: |
| --- | --- |
| 1. 21-70 years of age<br>2. Able to understand and provide an informed consent or have legally authorized representative consent to participate<br>3. Able to understand and comply with study procedures<br>4. Access to Wi-fi at home; and an iPhone for uploading console data and participating in COR Research App<br>5. Willing to milk blood onto dried blood spot cards.<br>6. Found to be generally healthy (free of diseases listed in exclusion criteria) | 1. Found to be medically unsuitable for participation in this study by the investigator or clinician<br>2. Pre-existing health condition or disability that would prevent participation in a program that includes changes to diet, exercise or sleep<br>3. Currently under the care of a physician for a medical condition<br>4. Currently on medically restricted diet<br>5. Immunocompromised and/or chronic terminal illnesses<br>6. Known heart dysrhythmia<br>7. Compromised circulation or peripheral vascular disease<br>8. Type 2 Diabetes<br>9. Pregnant<br>10. Reported heart attack (myocardial infarction), stroke/transient ischemic attack, or major surgery in the last two years.<br>11. Unwilling to participate in COR Programs<br>12. Aversion to fingerstick<br>13. Live outside the US |

